# Radical versus partial nephrectomy and survival from stage T1 renal cell carcinoma in three prospective cohorts

**DOI:** 10.1101/2021.10.17.21265105

**Authors:** Christopher M. Sauer, Sarah C. Markt, Lorelei A. Mucci, Alejandro Sanchez, Steven L. Chang, Rebecca E. Graff, Mark A. Preston

## Abstract

**Background:** Whether or not a survival difference exists between radical and partial nephrectomy for stage T1 renal cell carcinoma (RCC) is controversial. We therefore aimed to evaluate cancer-specific, other cause, and overall survival among patients undergoing radical or partial nephrectomy for stage pT1 RCC.

**Materials and methods:** We identified 330 participants with pT1a-b RCC diagnosed between 2000-2015 in three prospective cohort studies and compared treatment with radical nephrectomy (N=196) versus partial nephrectomy (N=134). The primary outcome was overall survival. Secondary outcomes were other-cause and cancer-specific mortality. Kaplan-Meier plots were used to visualize overall survival for the two treatment groups. Cox proportional hazards regression was utilized to compare outcomes between groups, and Fine and Gray competing risks regression was used to compare cancer-specific and other cause mortality between groups. Multivariable models adjusted for age, tumor size, sex, year of diagnosis, body mass index, history of smoking, history of hypertension, surgical technique, and pathological differentiation.

**Results:** During a median follow-up of eight years, overall survival was 84%. We did not detect a statistically significant difference in overall survival between partial and radical nephrectomy (Hazard Ratio (HR) = 0.84, 95% Confidence Interval: 0.40-1.78). There was no significant difference in cause-specific or other cause mortality between groups. This study had 80% power to detect an HR ≥2.20.

**Conclusions:** These results did not suggest a difference in long-term survival outcomes between radical and partial nephrectomy.

## Introduction

Survival rates of localized kidney cancer in the United States are high, with approximately 93% of patients surviving 5 years ^1^. Historically, localized kidney cancer (T1-T2N0M0) has been treated by radical nephrectomy – complete removal of the affected kidney – resulting in good local disease control ^2^. The procedure, however, is associated with two major disadvantages. First, radical nephrectomy can have serious side effects, including chronic kidney disease and poor functional outcomes relating to potential long-term adverse effects of nephrectomy, such as anemia or in rare cases, dialysis ^3^. Second, radical nephrectomy might over-treat small kidney cancers that could be resected by a more targeted approach. As a result, surgeons have sought kidney-sparing surgical approaches, and partial nephrectomy has emerged as an alternative.

Guidelines, including those from the American Urological Association, now recommend nephron-sparing rather than radical surgical interventions for stage cT1a renal masses^4^. This consensus was reached based on non-inferior oncologic outcomes and lower rates of chronic kidney disease ^5^. Meanwhile, advantages of radical nephrectomy include shorter surgery and anesthesia time, and less complications (e.g., urine leak) ^5^.

Numerous studies have contrasted radical and partial nephrectomy with respect to outcomes, including overall survival, cancer specific survival, rates of renal failure and cardiovascular comorbidities ^6-9^. A 2010 randomized controlled trial (EORTC 30904) comparing radical with partial nephrectomy for kidney cancers ≤5cm found a trend towards better overall survival in the radical nephrectomy group compared to the partial nephrectomy group (10-year overall survival: 81.1% for radical vs. 75.7% for partial nephrectomy). However, these findings were no longer statistically significant when restricting to renal cell carcinoma (RCC), thus excluding benign renal masses, as *a priori* defined in the study protocol ^10^.

In contrast, a systematic review of observational studies ^5^ found partial nephrectomy to have superior overall survival rates (Pooled effect estimate: 0.81, 95% confidence interval, CI: 0.74, 0.89). The difference in the cancer specific survival trended in the same direction but was not statistically significant (Pooled effect estimate 0.85, 95% CI: 0.73, 1.01). An important limitation of prior observational studies is incomplete adjustment for confounding factors: For instance, patients with a history of hypertension or smoking could be more likely to receive partial nephrectomy and could be more likely to die. As a result, analyses not adjusting for these factors are at risk of residual confounding, likely favoring partial over radical nephrectomy ^7-9, 11^. In addition, many studies suffer from inadequate length of follow-up ^5^, which limits conclusions regarding long-term survival.

We aimed to address prior limitations and study long-term survival differences between partial and radical nephrectomy using three nationwide prospective cohort studies.

## Materials and Methods

### Study population

These analyses were conducted within three prospective cohort studies: the Nurses’ Health Study (NHS, established 1976; 121,700 female nurses aged 30-55 years), Nurses’ Health Study II (NHSII, established 1989; 116,429 female nurses aged 25-42 years) and Health Professionals Follow-up Study (HPFS, established 1986; 51,529 male health professionals aged 40-75 years). The cohorts have previously been described ^12^. Biennially, questionnaires are sent to participants to ascertain information on general health, recent diagnoses, lifestyle factors, diet (every four years), physical activity, and medication use.

The study protocol was approved by the institutional review boards of the Brigham and Women’s Hospital (2005P000173/BWH) and Harvard T.H. Chan School of Public Health, and those of participating registries as required.

### Identification of cases and treatment modalities

Kidney cancer cases were initially identified by self-report or participants’ next of kin on biennial questionnaires and confirmed by medical record and pathology report review by two of the authors (MAP, AS). Information on TNM stage, differentiation, and Fuhrman grade was extracted from the original hospital’s pathology reports, and information on treatment modalities and timing was collected from medical records. Analyses were restricted to pT1 cases of clear cell, papillary, chromophobe, or unclassified RCC who underwent radical or partial nephrectomy between 2000 and 2015. This was done to (a) minimize variance in prognosis due to pathological subtype, and tumor stage and (b) reduce any potential introduction of bias from time-varying exposures. To validate the generalizability of findings, sensitivity analyses were performed including all cases (diagnosed 1977-2015) and, separately, including individuals diagnosed with stage T2N0M0 disease after 2000.

### Outcomes

Deaths in the cohorts are identified via reports from family members and linkage with the National Death Index, with greater than 98% completeness of follow-up ^13^. Cause of death, including from RCC, is assigned by an endpoints committee based on review of all available medical and autopsy reports.

### Statistical analysis

Cox proportional hazards models were fit to evaluate the association of surgery modality with all-cause mortality. Time to event was calculated from the month of diagnosis until the time of death or administrative censoring (December 2015). Kaplan-Meier plots were used to visualize survival for both groups ^14^. Fully adjusted models included age at diagnosis, tumor size (continuous, cm), sex (female / male), year of diagnosis (ordinal, 2000 to 2015), body mass index (BMI; categorical, <18.5, 18.5-24.9, 25.0-29.9, >30 kg/m^2^), history of smoking (never / ever), history of hypertension (yes / no), history of diabetes (yes / no), surgical technique (open / minimally invasive), and pathological differentiation (well-moderate / poor). Schoenfeld’s global test was used to evaluate the proportional hazards assumption.

In a sensitivity analysis, we performed competing risks regression to study differences in death from kidney cancer and other causes of death ^15^ according to surgery modality. In two separate models, the frequency of onset of *de novo* diabetes and hypertension as outcomes was compared between the surgical modality groups.

Predicted cumulative incidence curves were plotted using the Nelson-Aalen estimator. To study whether differential length of follow-up affected the results, we performed a sensitivity analysis in which we excluded all cases with less than 2 years of follow-up. Lastly, we tested whether there existed differences in the survival curves from diagnosis to death in the time intervals ranging from 0-4.9, 5-10, and >10 years.

All analyses were performed using *R* (version 3.4, http://www.R-project.org/) and relevant packages. All tests were two-sided and used an alpha-level of 0.05 to determine statistical significance.

## Results

Across all the cohorts, we identified 911 kidney cancer cases. Of these, 782 underwent partial or radical nephrectomy and 465 had pathological T1N0M0 stage. Since few cases underwent partial nephrectomy before 2000 due to the underling age structure of the cohorts, primary analyses were restricted to those who underwent surgery between 2000 and the end of follow-up in 2015 (N= 330). Of these, 196 underwent radical nephrectomy and 134 partial nephrectomy (Table 1). Most cases were identified in the NHS I (N=149), with the HPFS contributing 101 cases and the NHS II contributing 80 cases. Mean length of follow-up was higher in the radical nephrectomy group (103 months), as compared to the partial nephrectomy group (84 months). The proportion of participants undergoing radical nephrectomy was highest in the NHS, while partial nephrectomy was more common in the HPFS and NHS II. Of note, participants undergoing radical nephrectomy tended to have higher rates of hypertension and higher BMI.

**Table 1:** Baseline characteristics of study participants according to treatment modality. *mean (sd), NHS: Nurses’ Health Study, NHS II: Nurses’ Health Study II, HPFS: Health Professionals Follow-up Study. ^&^ Not adjusted for length of follow-up

| Variable | Level | Radical<br>nephrectomy | Partial<br>nephrectomy |
| --- | --- | --- | --- |
| Number of participants |  | 196 | 134 |
| Cohort (%) | HPFS | 53 (27.0) | 48 (35.8) |
|  | NHS | 101 (51.5) | 48 (35.8) |
|  | NHS II | 42 (21.4) | 38 (28.4) |
| Year of diagnosis | 2000-2004 | 71 (36.2) | 39 (29.1) |
|  | 2005-2009 | 75 (38.3) | 35 (26.1) |
|  | 2010-2015 | 50 (25.5) | 60 (44.8) |
| Follow-up, months* |  | 103.5 (46.6) | 83.5 (46.0) |
| Age at diagnosis, years* |  | 67.9 | 66.4 |
| Male (%) |  | 53 (27.0) | 48 (35.8) |
| History of smoking (%) |  | 100 (51.0) | 66 (49.3) |
| Hypertension (%) |  | 118 (60.2) | 70 (52.2) |
| Diabetes (%) |  | 32 (16.8) | 16 (13.1) |
| Body Mass Index (%) | <18.5 | 2 (1.0) | 0 (0.0) |
|  | 18.5- 24.9 | 51 (26.0) | 35 (26.1) |
|  | 25- 29.9 | 73 (37.2) | 66 (49.3) |
|  | >30 | 70 (35.7) | 33 (24.6) |
| Type RCC (%) | Clear cell | 149 (76.0) | 95 (70.9) |
|  | Papillary | 21 (10.7) | 24 (17.9) |
|  | Chromophobe | 16 (8.2) | 13 (9.7) |
|  | RCC unclassified | 10 (5.1) | 2 (1.5) |
| Pathological<br>Differentiation (%) | Good/ intermediate | 145 (74.0) | 92 (68.7) |
|  | Poor | 34 (17.3) | 27 (20.1) |
|  | Missing | 17 (8.7) | 15 (11.2) |
| Reason for diagnosis (%) | Hematuria | 16 (8.2) | 14 (10.4) |
|  | Flank pain | 12 (6.1) | 6 (4.5) |
|  | Incidental | 114 (58.2) | 85 (63.4) |
|  | Systemic | 2 (1.0) | 0 (0.0) |
|  | missing | 52 (26.5) | 29 (21.6) |
| Tumor stage | T1a (<4cm) | 91 (46.4) | 104 (77.6) |
|  | T1b (≥4cm) | 105 (53.6) | 30 (22.4) |
| Open surgery (%) |  | 67 (47.9) | 49 (52.7) |
| Death<sup>&</sup> (%) | Due to RCC | 4 (2.0) | 2 (1.5) |
|  | Due to other<br>causes | 34 (17.3) | 13 (9.7) |

Patients treated with partial nephrectomy more frequently had T1a stage tumors (77.6%) than patients treated with radical nephrectomy (46.4%). A total of 53 participants died from any cause during a mean follow-up time of 95 months, with a similar proportion dying from RCC in both groups (2.0% versus 1.5%). Crude mortality from other causes of death was higher in the radical nephrectomy group (17.3%) compared to the partial nephrectomy group (9.7%). In both groups, among patients with recorded causes of death, the most frequent one was cardiovascular disease (radical nephrectomy: 7, partial nephrectomy: 3).

Treatment allocation to radical or partial nephrectomy varied with time (**Figure 1**). RCC cases diagnosed before 2007/2008 were more frequently treated with radical nephrectomy compared with more recent cases (p=0.002).

**Figure 1:**
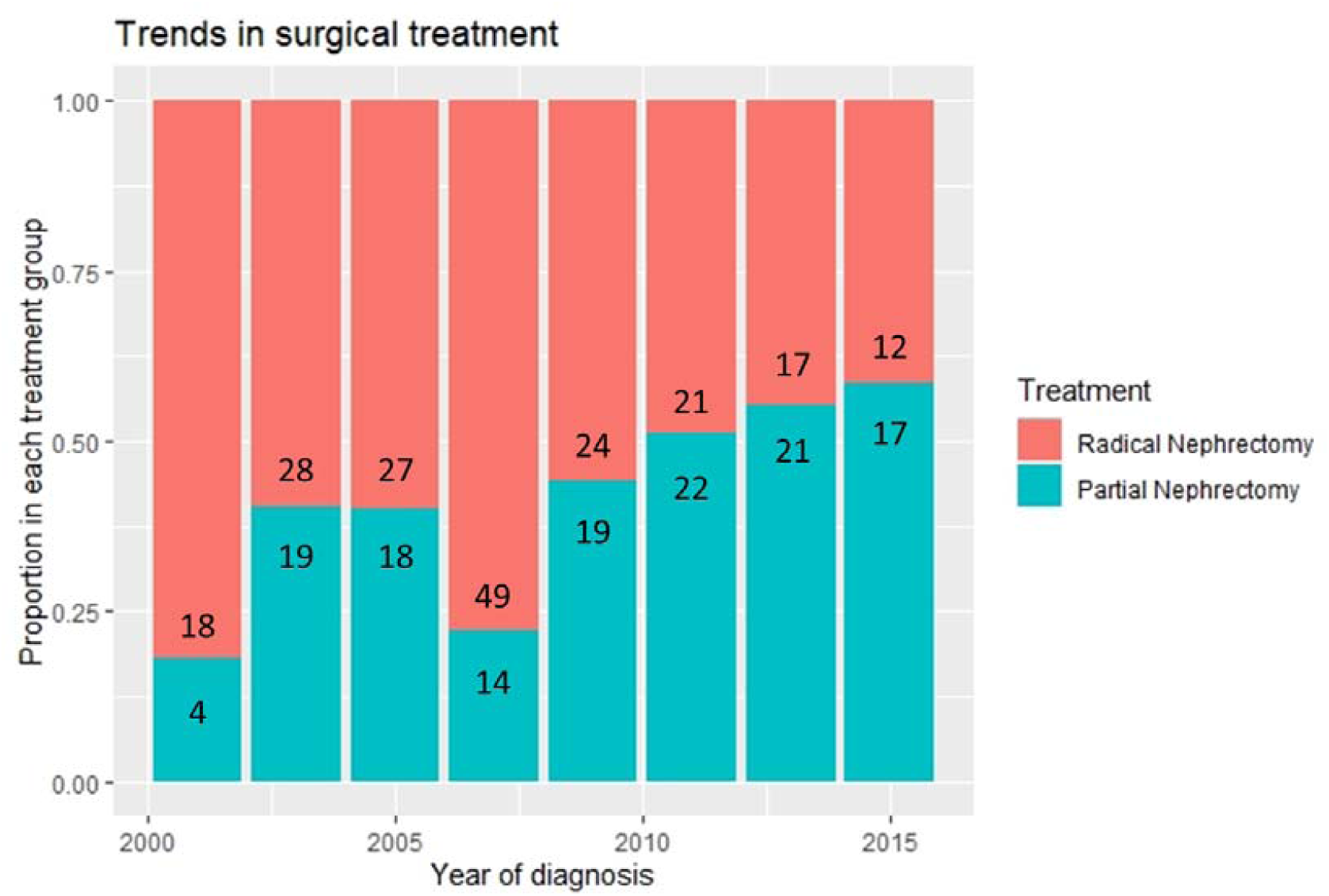
Proportion of individuals undergoing radical or partial nephrectomy over time. Numbers within the bars represent absolute case counts for each treatment and period.

In the univariate analysis, we did not find a statistically significant difference in overall survival between participants undergoing radical and partial nephrectomy (HR=0.82, 95%CI: 0.45-1.50). A very similar estimate was obtained after adjustment for age and tumor size, (HR=0.84, 95%CI: 0.44-1.60). Even after full adjustment for covariates, we did not find a statistically significant difference (HR=0.84, 95%CI: 0.40-1.78, Table 2).

**Table 2:** Clinical, demographic, and epidemiological factors associated with all cause-mortality. Only age and presence of diabetes at diagnosis were significantly associated with all-cause mortality. There was no difference in the hazard of death between the radical and partial nephrectomy groups.

| **Variable** | **Hazard Ratio** | **95% Confidence Interval** | **p-value** |
| --- | --- | --- | --- |
| Partial Nephrectomy | 0.84 | 0.40-1.78 | 0.65 |
| Year of diagnosis | 0.91 | 0.79-1.03 | 0.14 |
| Age at diagnosis | 1.10 | 1.05-1.15 | **<0.001** |
| Male sex | 1.54 | 0.81-2.96 | 0.19 |
| History of Smoking | 1.57 | 0.89-2.77 | 0.12 |
| History of Hypertension | 1.22 | 0.61-2.44 | 0.58 |
| History of Diabetes | 2.81 | 1.40-5.67 | **0.004** |
| BMI 25-29.9 | 0.75 | 0.36-1.57 | 0.45 |
| BMI >30 | 0.94 | 0.40-2.17 | 0.88 |
| Tumor size, in cm | 0.89 | 0.72-1.11 | 0.31 |
| Poor differentiation | 1.45 | 0.71-2.97 | 0.31 |
| Open surgery | 1.06 | 0.47-2.37 | 0.89 |

There was no evidence for a difference in predicted OS between radical and partial nephrectomy, with the survival curves being very similar (Figure 2). No trend towards differential survival probabilities after long-term follow-up (>10 years) was found.

**Figure 2:**
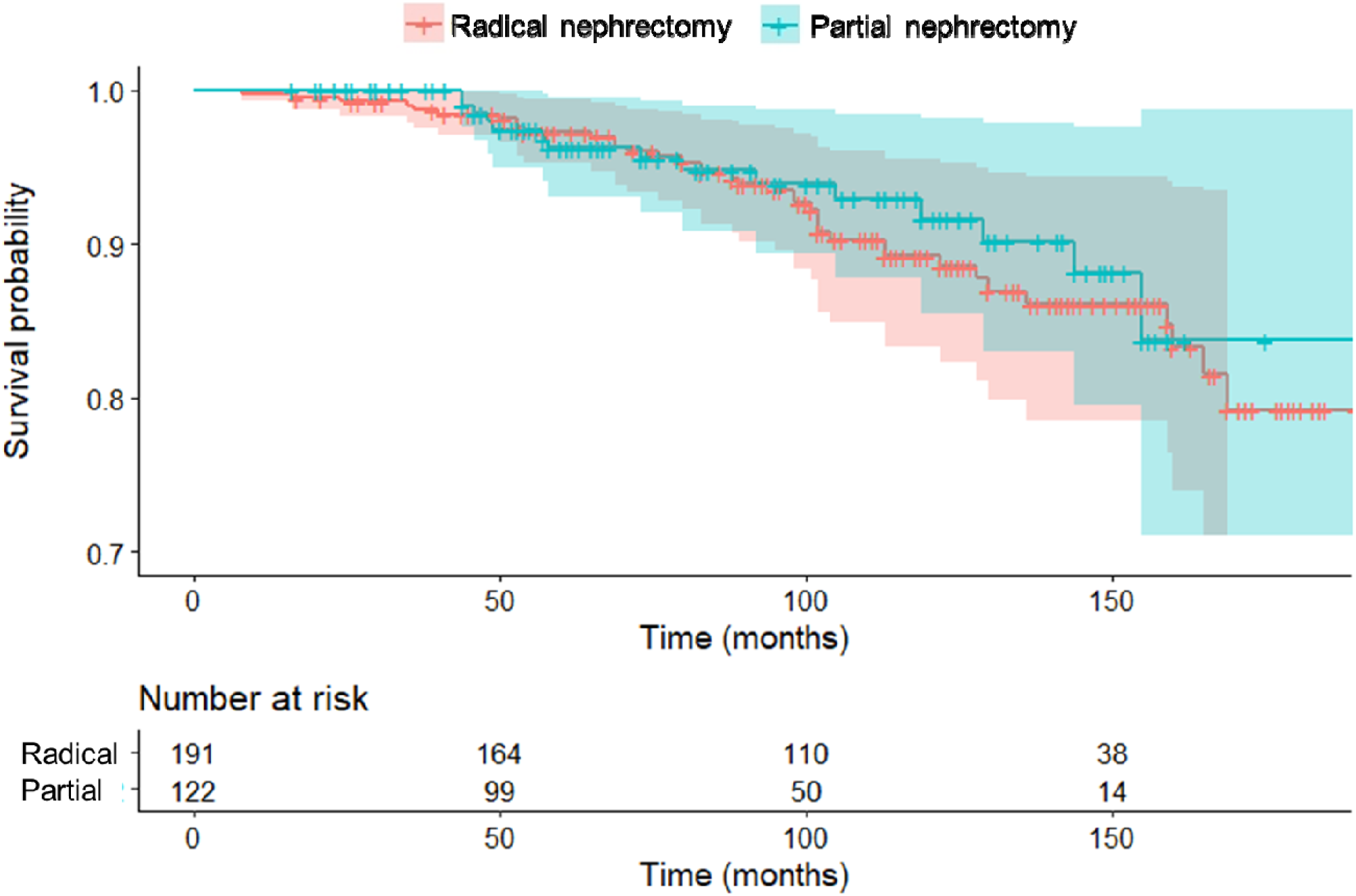
Predicted over survival from the fully adjusted Cox proportional hazards model. Shaded areas represent 95% confidence intervals.

Based on competing risks regression, no significant difference between the surgical techniques was found with respect to death from other causes (HR=0.80, 95%CI: 0.42-1.51, p=0.49). To study potential mechanisms related to differential mortality between the surgical techniques we compared the odds ratios of new onset hypertension and diabetes; however, we did not find a difference in the onset of *de novo* hypertension (OR=1.07, 95%CI: 0.58-1.94, p=0.89) or *de novo* diabetes (OR=2.01, 95%CI: 0.33-13.96).

Sensitivity analyses, including the inclusion of cases diagnosed before 2000 (N= 465), and, separately, including pathological stage T2 cases (N= 380) returned materially similar results. In a separate analysis, no statistically significant difference between the treatment modalities for <5, 5-10, and >10 years of follow-up was found (all p>0.20).

## Discussion

In this prospective analysis of 330 cases with pT1 RCC, we did not find a difference in overall or cancer-specific survival of individuals undergoing radical nephrectomy compared to those undergoing partial nephrectomy. The results do not support earlier findings that reported a survival benefit of patients undergoing partial nephrectomy ^6, 16-18^. A potential explanation could be that many earlier studies did not fully adjust for the most common factors that influence the choice between radical and partial nephrectomy (e.g., obesity, hypertension, and a history of smoking). Obese participants are more likely to undergo radical nephrectomy and obesity is also a strong risk factor for overall death ^19^. Similarly, history of hypertension, and to a lesser extent history of diabetes or smoking reflect poorer overall health and are more prevalent among participants undergoing radical nephrectomy (Table 2).

Interestingly, we observed that the inclusion of additional confounders in the Cox proportional hazards model had little effect on the treatment estimates. Compared with other studies, including within the Surveillance, Epidemiology and End Results (SEER) database (2004–2014) ^8^ which adjusted for fewer confounders, our findings suggest no difference in outcomes. Of note, the findings presented in this paper were robust when performing sensitivity analyses, such as including cases before 2000, or T2 tumors.

Furthermore, many prior studies do not perform subgroup analyses but rather compare outcomes for all types of kidney cancers. ^20^. Since kidney cancer histologic subtypes are clinically and genomically distinct entities ^21^ with divergent prognoses ^22^, analyses should separate results for the most common types of RCC from spindle-cell and collecting duct tumors, which have different prognosis. Of note, we did not find a difference in survival between the surgical modalities in both the crude and the fully adjusted analysis, which could potentially be due to less pronounced differences in patient characteristics at baseline.

In the before mentioned phase III non-inferiority EORTC30904 randomized trial ^10^, there was also no statistically relevant difference in 10 years overall survival for RCCs between the radical and partial nephrectomy arm (HR 1.34, p=0.17).

The participants included in this study were derived from nation-wide cohort studies and their characteristics were very similar to national average RCC patients with regards to gender, age at diagnosis, and tumor size ^23^. Furthermore, survival rates were similar to national averages and the European trial ^1, 10^. To the best of our knowledge, the mean follow-up of 8 years after cancer diagnosis is the longest in the published literature. Additional important advantages of our analysis include its prospective nature, high rates of follow-up over more than 30 years, detailed availability of clinical and baseline characteristics, ascertainment of disease status through review of pathology reports, and adjustment for important confounders. We thus believe that the results of this study are generalizable to most U.S. settings.

The main limitation is the low event rate, which limited the statistical power to detect a difference in overall survival between treatments. With the observed number of events, we had approximately 80% power to detect a HR≥2.2. Furthermore, due to the low number of participants dying from RCC, it was infeasible to demonstrate a statistically significant association between treatment and RCC-specific death. While we were able to adjust for the most important confounders of treatment allocation and survival, we were unable to include information on anatomical location and tumor complexity, e.g. using the renal nephrometry score ^24^, or additional patient characteristics relating to pre-surgical comorbidities. While data on new onset of hypertension was available, we did not have access to pre- and post-surgery kidney function data, thus were unable to analyze potential differences in estimated glomerular filtration rates (eGFR) between the groups. Lastly, we would like to highlight that partial nephrectomy has only lately become widely used, thereby rendering the median length of follow-up longer in the radical nephrectomy group. By using time-to-event approaches and accounting for potential competing risks, we mitigated these differences, though it should be noted that few participants were at risk of death in the partial nephrectomy group after more than 10 years of follow-up.

For routine clinical practice, we conclude based on this study that there is insufficient evidence that either surgical approach is superior with regards to overall survival. We therefore suggest an individualized approach for each patient, taking into account surgical feasibility, surgeons’ experience, patient’s risk profile and preference.

## Conclusions

In this prospective cohort study, we showed no difference in long-term survival between participants undergoing radical versus partial nephrectomy after adjustment for confounders. Future studies could help in the decision-making process by including kidney function measures as well as quality of life outcomes.

## Data Availability

Data used for this analysis is not publicly available but restricted access can be discussed upon reasonable request.

## Acknowledgements

This study relied on data from the NHS I, NHS II and HPFS, which received grant support from the National Institutes of Health (grant numbers UM1 CA186107, P01 CA87969, U01 CA176726, U01 CA 167552). The content is solely the responsibility of the authors and does not necessarily represent the official views of the National Institutes of Health.

We would like to thank the following state cancer registries for their help: AL, AZ, AR, CA, CO, CT, DE, FL, GA, ID, IL, IN, IA, KY, LA, ME, MD, MA, MI, NE, NH, NJ, NY, NC, ND, OH, OK, OR, PA, RI, SC, TN, TX, VA, WA, WY. The authors assume full responsibility for analyses and interpretation of these data.

We would like to thank the Channing Division of Network Medicine, Department of Medicine, Brigham and Women’s Hospital for their support reviewing this manuscript.

